# Association of Lacunes with Risk Factors, Cognition, and Atrophy: The Multi-Ethnic Study of Atherosclerosis (MESA)

**DOI:** 10.64898/2026.07.02.26357192

**Authors:** David H. Wang, Faiza Azhar, Niyas Shamsudeen, Jose Gutierrez, Sokratis Charisis, Sachintha Ransara Brandigampala, Kyle C. Kern, Tanweer Rashid, Paul Jensen, Ilya Nasrallah, Jeffrey B. Ware, Kevin Hiatt, Jordan Tanley, R. Nick Bryan, WT Longstreth, Lenore Launer, Sudha Seshadri, Timothy M. Hughes, Susan R. Heckbert, Mohamad Habes

**Affiliations:** Neuroimage Analytics Laboratory (NAL) and the Biggs Institute Neuroimaging Core (BINC), Glenn Biggs Institute for Alzheimer’s and Neurodegenerative Diseases, University of Texas Health Science Center San Antonio, San Antonio, Texas; Department of Neurology, University of Miami, Miami, Florida; Department of Neurology, Massachusetts General Hospital, Boston, Massachusetts; Harvard Medical School, Boston, Massachusetts; Department of Neurology, UCLA David Geffen School of Medicine and West LA Veterans Affairs Medical Center, Los Angeles, California; Department of Epidemiology, University of Washington, Seattle, Washington; AI2D Center for AI and Data Science for Integrated Diagnostics, and Center for Biomedical Image Computing and Analytics, University of Pennsylvania, Philadelphia; Department of Radiology, Perelman School of Medicine, University of Pennsylvania, Philadelphia; Department of Radiology, Wake Forest School of Medicine, Winston-Salem, North Carolina; Department of Internal Medicine, Wake Forest University School of Medicine, Winston-Salem, North Carolina; Department of Neurology, University of Washington, Seattle, Washington; Intramural Research Program, Laboratory of Epidemiology and Population Sciences, National Institute on Aging, National Institutes of Health, Bethesda, Maryland; Glenn Biggs Institute for Alzheimer’s and Neurodegenerative Diseases, University of Texas Health Science Center San Antonio, San Antonio, Texas

**Keywords:** lacunes, cerebral small vessel disease, magnetic resonance imaging, deep learning, cognitive decline, cardiovascular risk factors

## Abstract

**Background:** Lacunes are 3-15 mm cavities originating from small perforating artery disease and are a hallmark of cerebral small vessel disease (cSVD). Prior prevalence studies relied on manual rating, which is prone to inter-rater variability and cannot quantify volume. We applied deep learning to quantify lacunes in a diverse community-based cohort.

**Methods:** In this cross-sectional analysis of 1,038 Multi-Ethnic Study of Atherosclerosis (MESA) participants, with longitudinal cognitive follow-up, we quantified lacunes, confirmed them with a trained rater, and classified them as deep or lobar. Regression models examined associations of lacunes with cardiovascular risk factors, other small vessel disease lesions, brain atrophy, and cognition. Structural equation modeling evaluated whether deep lacune burden mediated associations of age or Framingham All-Cardiovascular Disease (CVD) Risk Score with atrophy and cognition.

**Results:** Overall, 182 participants (17.5%) had at least one lacune (deep: 9.2%; lobar: 9.6%). Hispanic participants had lower lacune burden than White participants. Age, Framingham All-CVD Risk Score, PREVENT 10-year Total CVD Risk Score, and hypertension were associated with overall and deep lacunes, while lobar lacunes showed no associations. Deep lacunes were associated with white matter hyperintensities, enlarged perivascular spaces, and cerebral microbleeds, as well as SPARE-BA, SPARE-AD, and cortical and hippocampal atrophy. Deep lacunes were cross-sectionally associated with decreased global cognition and language/semantic performance, and longitudinally with accelerated executive function decline independent of count. Deep lacune burden significantly mediated associations of age and Framingham All-CVD Risk Score with SPARE-AD, SPARE-BA, and global cognition.

**Conclusions:** In this multi-ethnic community-based cohort, deep lacune burden demonstrated stronger associations with vascular risk, other cerebral small vessel disease markers, brain atrophy, and longitudinal executive function decline than lobar burden. By providing a continuous measure of lesion volume, automated quantification captured information beyond lacune count.

## 1. Introduction

Lacunes are round or ovoid fluid-filled cavities between 3 and 15 mm, resembling cerebral spinal fluid (CSF). While lacunes can be caused by emboli, most result from diseases of the small deep perforating arteries or pathology in their parent vessel^1^. While lacunes historically referred to ischemic lesions on pathology affecting the deep brain structures^2^, the STandards for ReportIng Vascular changes on nEuroimaging (STRIVE) criteria define lacunes radiologically as lesions of the perforating artery territory^3^. But there is an increasing body of evidence that these frequently subclinical lesions^4–6^ have a disproportionate effect on brain health, since they are associated with increased risk of future stroke, gait impairment, dementia, and death^7^. Lacunes are primarily identified in-vivo through visual reads of fluid-attenuated inversion recovery (FLAIR) and T1-weighted (T1w) magnetic resonance imaging (MRI). According to the STRIVE criteria, lacunes are small and subcortical lesions that appear hypointense on FLAIR MRI with a characteristic hyperintense rim, hypointense on T1w MRI, and hyperintense on T2-weighted (T2w) MRI^3^. Visual identification is challenging as the hyperintense rim is not always present, and the cavitation in some instances may be incompletely suppressed on FLAIR, despite showing CSF-like qualities on T1w and T2w MRI^8^.

Most previously published studies of the prevalence of lacunes used manual rating scales, leveraging the visual inspection of FLAIR, T1w and T2w MRI to manually count the number of lesions that met the STRIVE criteria^4–6,9–21^. However, this approach is labor intensive, and manual reads suffer either from inter-rater variability or single-reader bias. Furthermore, manual reads are unable to quantify three-dimensional volume of lesions.

The Multi-Ethnic Study of Atherosclerosis (MESA) is a prospective, community-based cohort designed to investigate subclinical cardiovascular disease, making it well-suited for studying subclinical lacune pathophysiology. Much prior lacune research relied on stroke-based and cerebral amyloid angiopathy (CAA) cohorts^9,10,17–22^ and on monoracial or biracial cohorts^5,11–15,17,21^, limiting generalizability to the subclinical lacunes that predominate in community settings. In contrast, MESA permits direct comparison of subclinical lacune burden across White, Black, Hispanic, and Chinese American participants within a single population-based sample.

In this paper, we leveraged our validated deep learning framework for small vessel disease quantification^23^ to study lacunes in the MESA cohort. We examined the associations of lacune burden with demographics, cardiovascular risk factors, other lesions of cerebral small vessel disease and gray matter atrophy patterns. We finally evaluated whether deep lacune burden statistically mediated the association between age as well as the Framingham All-Cardiovascular Disease (CVD) Risk Score^24^ with brain atrophy patterns and cognitive decline.

## 2. Methods

### 2.1. Participants

MESA enrolled 6,814 participants aged 45-84 years at baseline across 6 field centers. Individuals with clinically apparent CVD were excluded, defined as a prior history of myocardial infarction, angina, stroke, heart failure, atrial fibrillation, or cardiac surgery^25^. MESA has conducted 6 follow-up examinations since its baseline examination in 2000-2002. At the 2016-2018 exam, 1,942 participants were invited to participate in the MESA Atrial Fibrillation ancillary study, of whom 1,062 underwent brain MRI. These participants did not have atrial fibrillation at baseline, but were monitored for the development of atrial fibrillation^26^. Each study site obtained institutional review board approval, and all participants provided written informed consent.

### 2.2. Demographics, Vascular Risk Factors, and Genetics

Demographic data were collected at the MESA baseline examination, including self-reported age, sex, and race/ethnicity amongst categories predefined by MESA investigators^25^. At the 2016-2018 exam, vascular risk factors were collected, including smoking status, medication use, and lipid panel. Blood pressure was calculated from the mean of the last two of three seated measurements and height and weight were measured. Hypertension was defined as systolic blood pressure ≥140 mmHg or diastolic blood pressure ≥90 mmHg, or use of antihypertensive medication. Fasting blood samples were obtained for blood glucose and hemoglobin A1c (HbA1c). Diabetes was defined as HbA1c ≥6.5%, fasting glucose ≥126 mg/dL, or use of diabetes medications. The Framingham All-CVD Risk Score (coronary heart disease, stroke, peripheral artery disease, or heart failure) was calculated using methodology described elsewhere^24^. Cardiovascular risk was also calculated using the American Heart Association’s Predicting Risk of cardiovascular disease EVENTs (PREVENT) equations^27^, implemented in R via the preventr package (v0.11.0). Because MESA’s bundled eGFR variable uses the older race-inclusive equation, eGFR was recomputed using the 2021 race-free CKD-EPI creatinine equation^28^ to match the PREVENT specification. APOE isoforms were estimated from single nucleotide polymorphisms rs429358 and rs7412^29^. History of stroke or transient ischemic attack are presented in Table S1. Distribution of risk factors are presented in Table S2.

### 2.3. Brain MRI Acquisition, Scanning Protocol, and Quality Control

Brain MRI acquisition for the MESA Atrial Fibrillation ancillary study was performed during 2018-2019 in conjunction with exam 6 on 3-Tesla Siemens scanners. Structural sequences included 1-mm isotropic sagittal three-dimensional T1w, T2w, and FLAIR imaging, as well as axial two-dimensional 30-direction echo-planar diffusion tensor imaging (DTI). The complete brain MRI protocol has been described elsewhere^30^.

All FLAIR, T2w, and T1w images were visually inspected for distortions and motion artifacts, and participants with poor quality scans were excluded from the analytic sample.

### 2.4. Cognitive Assessment

MESA measured the Cognitive Abilities Screening Instrument (CASI), Digit Symbol Coding (DSC), and Digit Span (DS) at 2016-2019, 2019-2021, and 2022-2024 exams at all sites and the comprehensive neuropsychological battery (Uniform Data Set version 3 [UDSv3], administered in English, Spanish, Mandarin, and Cantonese) in 2016-2019 (limited to Wake Forest and Johns Hopkins sites) and at all MESA sites in 2019-2021 and 2022-2024, yielding domain scores for immediate memory, delayed memory, language/semantic fluency, phonemic fluency, attention/processing speed, executive function, and visuospatial functioning^31^. Individual test scores were normalized using published consensus norms^32,33^ or MESA-derived norms, and domain scores were calculated as the average of constituent normalized tests. These cognitive domains were normalized for age, education, race/ethnicity, sex, and language of administration^31^. A global cognitive score was calculated through the average of the z-scores of CASI, DSC, and DS. The complete cognitive protocol has been described elsewhere^34,35^. Cognitive assessment completeness is shown in Figure S1.

### 2.5. Manual Annotation of Lacune Ground Truth

Annotation of lacunes was based on the STRIVE criteria^3^. All segmentations were done on the ITK-SNAP software. The annotation protocol is presented in Figure S2.

Ground truth criteria were established based on consensus of two independent readers. Readers achieved a mean lesion-wide κ of 0.771.

### 2.6. Model Architecture and Training

We modified our previous U-Net architecture^23^ for automated lacune segmentation from FLAIR, T2w, and T1w imaging. The network incorporated ResNet blocks in place of standard convolutional layers, LeakyReLU activations, and instance normalization. The model was trained on 47 ground truth cases with at least one lacune. The model was trained with a depth of 5, an initial filter size of 32, a batch size of 32, and a learning rate of 0.0001. Model performance was evaluated with 5-fold cross validation. Demographic of the participants whose scans were used for ground truth are presented in Table S3.

### 2.7. Lacune Segmentation, Mapping, and Quality Control

We segmented lacunes from FLAIR, T2w, and T1w imaging using our trained deep learning model. Regional lacune burden was quantified using the MUlti-atlas region Segmentation utilizing Ensembles (MUSE) atlas^36^. Lesions in multiple regions were assigned a primary region by greatest overlap. Because precision was modest, all segmented lesions underwent visual quality control by a trained rater before inclusion in the analytic dataset, following a procedure consistent with the manual annotation protocol described above. Segmentation masks were also adjusted by trained rater if necessary. The standard operating procedure (SOP) for our developed QC tool is presented in Supplemental Methods. Downstream analyses used reader-confirmed lacune masks rather than raw model output. Lacunes were then mapped to deep and lobar lacunes based on Microbleed Anatomical Rating Scale (MARS)^37^-derived MUSE mapping. Brainstem lacunes were excluded due to lack of adequate sample. An example of deep and lobar lacunes can be found in Figure 1A. MUSE to MARS mapping is presented in Table S4.

**Figure 1.**
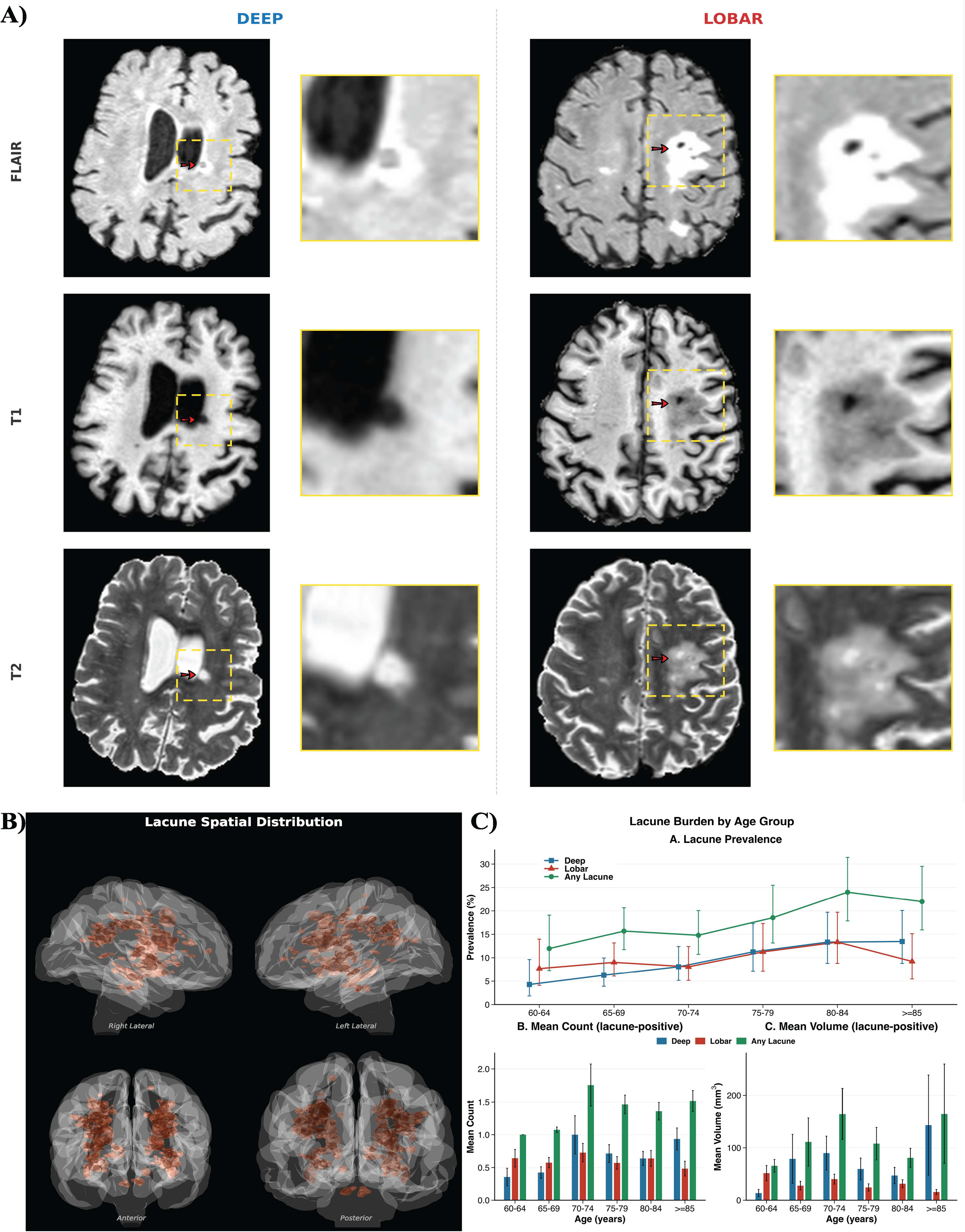
Representative Deep and Lobar Lacunes on Multimodal MRI, Lacune Burden by Age Group, and Spatial Distribution. Panel A shows representative examples of a deep lacune and a lobar lacune on axial FLAIR, T1-weighted, and T2-weighted MRI. Red arrows indicate the lacune; yellow insets show magnified views. On FLAIR, lacunes appear as hypointensities with a hyperintense rim. On T1-weighted imaging, lacunes are hypointense, and on T2-weighted imaging, hyperintense, consistent with CSF-like cavitation per STRIVE criteria. Panel B shows lacune prevalence, mean count, and mean volume stratified by 5-year age bands. Error bars indicate 95% CIs (prevalence) or standard errors (count, volume). Mean count and volume were calculated amongst participants with at least one lacune. Panel C shows the spatial distribution of all segmented lacunes projected onto a glass brain in right lateral, left lateral, anterior, and posterior views. Abbreviations: CI, confidence interval; CSF, cerebrospinal fluid; FLAIR, fluid-attenuated inversion recovery; MRI, magnetic resonance imaging; STRIVE, STandards for ReportIng Vascular changes on nEuroimaging.

### 2.8. Calculation of atrophy indices and segmentation of small vessel disease lesions

Spatial PAttern REcognition of Alzheimer’s Disease (SPARE-AD) and Spatial PAttern REcognition of Brain Aging (SPARE-BA) indices are support vector machine derived indices from T1-weighted MRI that quantify AD-related and BA-related atrophy. Regional volumetric and cortical thickness measures serve as input features, and each index produces a single continuous score per participant, where higher values indicate greater pathological burden. SPARE-AD was trained to distinguish AD from cognitively normal participants, capturing patterns of hippocampal and temporoparietal atrophy characteristic of AD^38^. SPARE-BA was trained to capture patterns of age-related structural change, providing a measure of brain aging beyond what is expected for a given chronological age^39^.

WMH volume was measured from inhomogeneity-corrected and co-registered FLAIR and T1w images using a validated deep learning-based segmentation method^38,40^ and grouped into periventricular and subcortical compartments^41^. Quantification of global, basal ganglia, and thalamus ePVS counts was performed from T2w, T1w, and FLAIR images using a validated deep learning-based model^23^. Cerebral microbleeds (CMB) counts were quantified from T2w images and susceptibility weighted imaging/quantitative susceptibility mapping using a validated deep learning-based model^42^. DTI-based measures of white matter microstructural integrity, including mean fractional anisotropy (FA) and mean diffusivity (MD), were calculated using automated DTI processing pipelines^43^.

### 2.9. Statistical Analysis

We performed a cross-sectional analysis of MESA, with a secondary longitudinal analysis of cognitive decline. 1038 participants had complete demographics, and quality-controlled MRI scans. Normality was assessed with Q-Q and kernel density plots. Lacune count and volume were cube-root transformed and subsequently z-scored. All continuous characteristics were z-scored prior to analysis so that effect estimates reflect a 1-SD change. CMB counts were cube-root normalized. WMH, ePVS, and SPARE-AD (shifted min plus 1) values were log-normalized. Participants were stratified into six 5-year age at MRI bands (60-64, 65-69, 70-74, 75-79, 80-84, ≥85). Prevalence of overall lacune, deep lacune, and lobar lacune was computed as the proportion of participants with at least one lesion of that subtype, with 95% confidence intervals calculated using the Wilson score method^44^. Among lacune-positive participants, mean lacune count and total volume were computed per age group to characterize lesion burden conditional on having at least one lacune.

Ordinary least squares (OLS) linear regression models examined direction of associations for cardiovascular risk factors with the dependent variables of lacune count and volume, reporting beta coefficients. Count and volume analysis included all participants, including those with no lacunes. Age, sex, Framingham All-CVD Risk Score, PREVENT 10yr Total CVD Risk Score, hypertension, high density lipoprotein (HDL) cholesterol, diabetes, current smoking status, race/ethnicity, and APOE ε4 carrier status were analyzed. Heteroscedasticity-consistent covariance matrix estimator type 3 (HC3) robust standard errors were applied to all regressions to account for heteroskedasticity. All models except Framingham All-CVD Risk Score and PREVENT 10yr Total CVD Risk Score models were adjusted for age, sex, race/ethnicity, MESA exam site, and educational attainment. Framingham All-CVD Risk Score and PREVENT 10yr Total CVD Risk Score analysis was only adjusted for race/ethnicity, MESA exam site, and educational attainment as age and sex are already considered in the risk scores^24,27^.

Race/ethnicity was modeled as a categorical predictor with White participants as the reference group. When a covariate served as the predictor of interest (such as age and sex), it was removed from the adjustment set.

Direction of association between lacune count and volume with the dependent variables of cognitive and neuroimaging outcomes were examined using OLS regression. Cognitive outcomes included the derived global cognitive score as well as all available cognitive domains including immediate memory, delayed memory, language/semantic fluency, phonemic fluency, attention/processing speed, executive function, and visuospatial functioning. Participants with only cognitive assessment in the 2022-2024 time frame were not included, due to large temporal difference between MRI and cognitive assessment. Neuroimaging outcomes included: SPARE-AD and SPARE-BA; Intracranial volume (ICV)-normalized cortical gray matter and hippocampal volumes; total, periventricular, and subcortical WMH; DTI FA and MD; total, basal ganglia, and thalamic ePVS counts; and total, deep, and lobar CMB counts. All models with neuroimaging outcomes and global cognition were adjusted for age, sex, race/ethnicity, study site, and education. WMH analyses were also adjusted for ICV. Global cognition analysis was also adjusted for language of test administration. Cognitive domain analyses were adjusted for the MESA site and the interval between MRI acquisition and cognitive testing. Age, sex, race/ethnicity, and education were omitted from cognitive domain models as cognitive domain scores were pre-normalized for these variables.

Linear mixed-effects (LME) models were used to assess longitudinal cognitive decline, with random slopes and intercepts for each participant. Baseline cognitive assessment was the earliest cognitive assessment for each participant. The association of lacunes with rate of cognitive decline was represented with the beta (β_Lacune×time_). To investigate the independent effect of lacune volume, lacune volume analysis was repeated with count as a covariate. As an additional sensitivity analysis, we repeated the LME models adjusting for global CMB count, WMH volume, and ePVS count to determine whether effect sizes were specific to lacunes independent of other cSVD lesions. The global cognitive score and all available cognitive domains were analyzed, with covariates mirroring cross-sectional analysis. Models were adjusted for the interval between MRI acquisition and the baseline cognitive assessment.

To evaluate whether deep lacune burden mediated the association between age and Framingham CVD risk score with brain atrophy and global cognition, structural equation modeling (SEM) was employed with a three-variable path model: Age or the Framingham All-CVD Risk Score was modeled as the independent variable, deep lacune burden as the mediator, and brain atrophy marker (SPARE-AD or SPARE-BA) or global cognition as the outcome. Deep lacune burden was operationalized as both count and volume in separate models, yielding twelve total models. Path coefficients were estimated using maximum likelihood estimation. The indirect effect was tested using bias-corrected bootstrap confidence intervals with 5,000 resamples. The proportion mediated was calculated as the indirect effect divided by the total effect. Covariates for age analysis included sex, education, site, and race/ethnicity. Covariates for Framingham analysis included education, site, and race/ethnicity. For all global cognition analysis, time between MRI and cognitive assessment as well as language of administration was added as a covariate.

Multiple comparisons were addressed using the Benjamini-Hochberg false discovery rate (FDR) procedure^45^. All analyses were FDR-corrected within each lacune classification (total, lobar, deep). A 2-sided *P* value ≤.05 was considered statistically significant. Any participants missing data for a particular analysis was excluded from that analysis.

## 3. Results

### 3.1. Participant Description, Lacune Distribution, and Model Performance

Participant characteristics are presented in Table 1. Due to the multiethnic nature of MESA, demographics were stratified by race/ethnicity. The analytic sample was racially and ethnically diverse, including Black, Hispanic, White, and Chinese American participants, with women comprising slightly over half of the cohort. Mean age was similar across racial/ethnic groups. During training and cross-validation on 47 ground truth cases, the lacune deep learning model achieved a sensitivity of 0.74 and precision of 0.41 in five-fold cross validation. All candidate lacunes were subsequently confirmed by a trained rater before analysis.

**Table 1.**
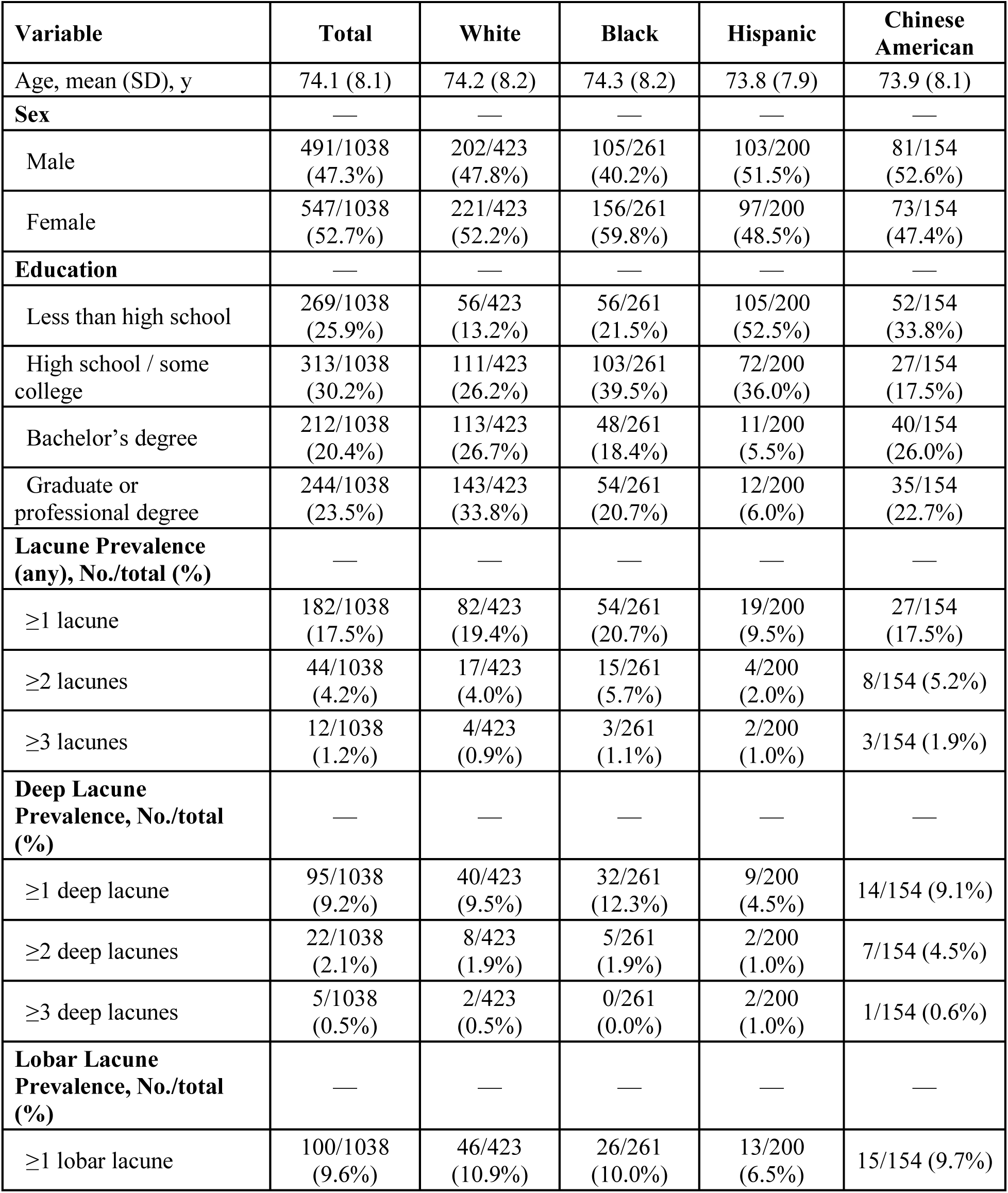
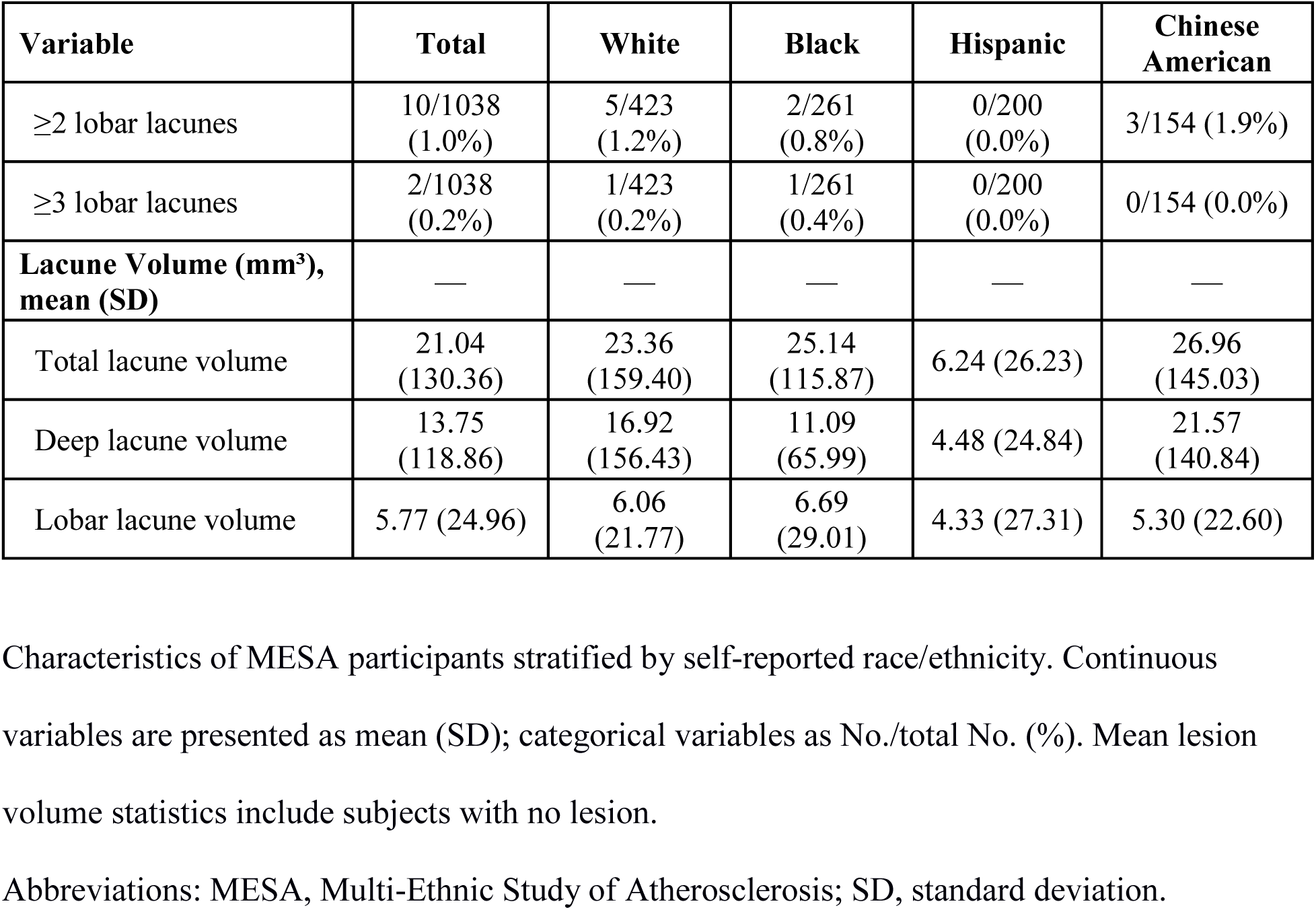
Demographic and Lacune Characteristics by Race/Ethnicity.

| **Variable** | **Total** | **White** | **Black** | **Hispanic** | **Chinese American** |
| --- | --- | --- | --- | --- | --- |
| Age, mean (SD), y | 74.1 (8.1) | 74.2 (8.2) | 74.3 (8.2) | 73.8 (7.9) | 73.9 (8.1) |
| **Sex** | — | — | — | — | — |
| Male | 491/1038<br>(47.3%) | 202/423<br>(47.8%) | 105/261<br>(40.2%) | 103/200<br>(51.5%) | 81/154<br>(52.6%) |
| Female | 547/1038<br>(52.7%) | 221/423<br>(52.2%) | 156/261<br>(59.8%) | 97/200<br>(48.5%) | 73/154<br>(47.4%) |
| **Education** | — | — | — | — | — |
| Less than high school | 269/1038<br>(25.9%) | 56/423<br>(13.2%) | 56/261<br>(21.5%) | 105/200<br>(52.5%) | 52/154<br>(33.8%) |
| High school / some college | 313/1038<br>(30.2%) | 111/423<br>(26.2%) | 103/261<br>(39.5%) | 72/200<br>(36.0%) | 27/154<br>(17.5%) |
| Bachelor's degree | 212/1038<br>(20.4%) | 113/423<br>(26.7%) | 48/261<br>(18.4%) | 11/200<br>(5.5%) | 40/154<br>(26.0%) |
| Graduate or professional degree | 244/1038<br>(23.5%) | 143/423<br>(33.8%) | 54/261<br>(20.7%) | 12/200<br>(6.0%) | 35/154<br>(22.7%) |
| **Lacune Prevalence (any), No./total (%)** | — | — | — | — | — |
| ≥1 lacune | 182/1038<br>(17.5%) | 82/423<br>(19.4%) | 54/261<br>(20.7%) | 19/200<br>(9.5%) | 27/154<br>(17.5%) |
| ≥2 lacunes | 44/1038<br>(4.2%) | 17/423<br>(4.0%) | 15/261<br>(5.7%) | 4/200<br>(2.0%) | 8/154 (5.2%) |
| ≥3 lacunes | 12/1038<br>(1.2%) | 4/423<br>(0.9%) | 3/261<br>(1.1%) | 2/200<br>(1.0%) | 3/154 (1.9%) |
| **Deep Lacune Prevalence, No./total (%)** | — | — | — | — | — |
| ≥1 deep lacune | 95/1038<br>(9.2%) | 40/423<br>(9.5%) | 32/261<br>(12.3%) | 9/200<br>(4.5%) | 14/154 (9.1%) |
| ≥2 deep lacunes | 22/1038<br>(2.1%) | 8/423<br>(1.9%) | 5/261<br>(1.9%) | 2/200<br>(1.0%) | 7/154 (4.5%) |
| ≥3 deep lacunes | 5/1038<br>(0.5%) | 2/423<br>(0.5%) | 0/261<br>(0.0%) | 2/200<br>(1.0%) | 1/154 (0.6%) |
| **Lobar Lacune Prevalence, No./total (%)** | — | — | — | — | — |
| ≥1 lobar lacune | 100/1038<br>(9.6%) | 46/423<br>(10.9%) | 26/261<br>(10.0%) | 13/200<br>(6.5%) | 15/154 (9.7%) |
| ≥2 lobar lacunes | 10/1038<br>(1.0%) | 5/423<br>(1.2%) | 2/261<br>(0.8%) | 0/200<br>(0.0%) | 3/154 (1.9%) |
| ≥3 lobar lacunes | 2/1038<br>(0.2%) | 1/423<br>(0.2%) | 1/261<br>(0.4%) | 0/200<br>(0.0%) | 0/154 (0.0%) |
| **Lacune Volume (mm<sup>3</sup>),**<br>**mean (SD)** | — | — | — | — | — |
| Total lacune volume | 21.04<br>(130.36) | 23.36<br>(159.40) | 25.14<br>(115.87) | 6.24 (26.23) | 26.96<br>(145.03) |
| Deep lacune volume | 13.75<br>(118.86) | 16.92<br>(156.43) | 11.09<br>(65.99) | 4.48 (24.84) | 21.57<br>(140.84) |
| Lobar lacune volume | 5.77 (24.96) | 6.06<br>(21.77) | 6.69<br>(29.01) | 4.33 (27.31) | 5.30 (22.60) |

Characteristics of MESA participants stratified by self-reported race/ethnicity. Continuous variables are presented as mean (SD); categorical variables as No./total No. (%). Mean lesion volume statistics include subjects with no lesion.
Abbreviations: MESA, Multi-Ethnic Study of Atherosclerosis; SD, standard deviation.

Overall, a subset of 182 participants had at least one lacune, with deep and lobar lacunes occurring at comparable frequencies. Lacune prevalence was highest in Black and White participants, and lowest in Hispanic participants. Deep lacune prevalence was highest in Black participants and lowest in Hispanic participants. Lobar lacunes were approximately equally prevalent in White, Black, and Chinese American participants, and lowest in Hispanic participants. Hispanic participants demonstrated the lowest burden across all lacune measures, including mean total lacune count and mean total lacune volume. Full demographics and lacune distribution can be found in Table 1. Spatial distribution of lacunes can be found in Figure 1B.

### 3.2. Lacune Prevalence by Age

Lacune prevalence increased progressively with age, approximately doubling from the youngest to oldest age strata. This trend was driven predominantly by deep lacunes, which rose approximately threefold from the youngest to oldest age strata, while lobar lacune prevalence remained relatively stable across age groups.

Among lacune-positive participants, mean lacune count and volume were highest in the 70-74 age group. Full distribution by age is shown in Figure 1C.

### 3.3. Association of Risk Factors with Lacune Burden

Older age was significantly associated with overall lacune count (β=0.11 [95% CI, 0.04-0.17]) and volume (β=0.09 [95% CI, 0.03-0.16]; Figure 2). Among subtypes, age was associated with deep lacune count (β=0.12 [95% CI, 0.06-0.19]) and volume (β=0.11 [95% CI, 0.05-0.18]), but not with lobar lacune measures. Sex was not significantly associated with any lacune measure.

**Figure 2.**
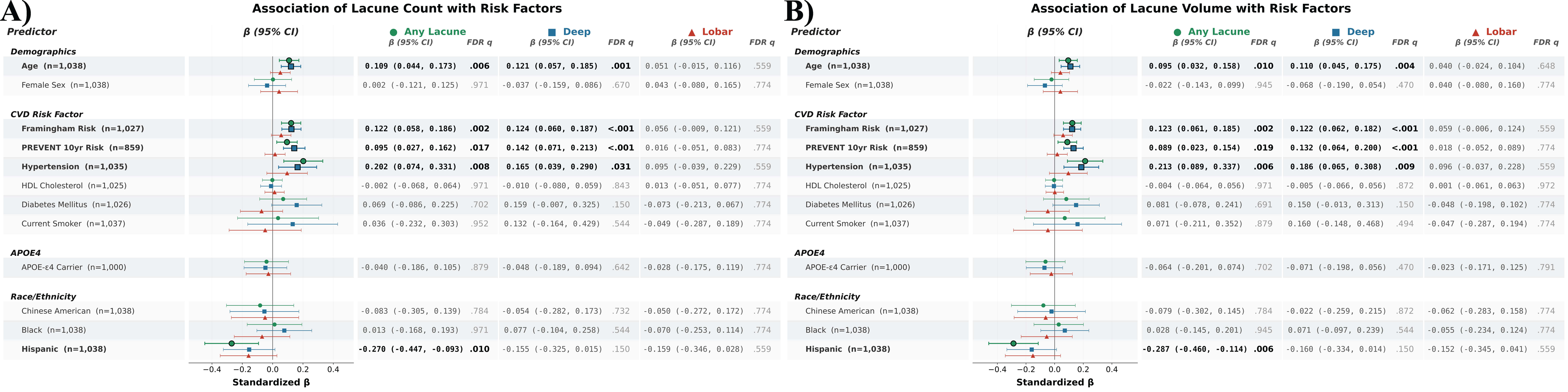
Association of Risk Factors With Lacune Burden. Forest plots showing associations of demographic, cardiovascular, genetic, and race/ethnicity predictors with lacune count (A) and lacune volume (B), stratified by lacune subtype (reference for race/ethnicity: White). Point estimates represent β coefficients with 95% CIs. Bold rows indicate FDR q < 0.05. Abbreviations: APOE ε4, apolipoprotein E ε4; CI, confidence interval; FDR, false discovery rate; HDL, high-density lipoprotein; SD, standard deviation.

The Framingham All-CVD Risk Score and the PREVENT 10yr Total CVD Risk Score were significantly associated with overall lacune count and volume (Framingham, β=0.12 [95% CI, 0.06-0.19] for both; PREVENT, β=0.09 [95% CI, 0.03-0.16] and β=0.09 [95% CI, 0.02-0.15]; Figure 2). Among subtypes, the Framingham All-CVD Risk Score (deep count, β=0.12 [95% CI, 0.06-0.19]; deep volume, β=0.12 [95% CI, 0.06-0.18]) and the PREVENT 10yr Total CVD Risk Score (deep count, β=0.14 [95% CI, 0.07-0.21]; deep volume, β=0.13 [95% CI, 0.06-0.20]) were associated with deep lacune count and volume, while neither lobar lacune volume nor count showed any associations.

Hypertension was significantly associated with overall lacune count (β=0.20 [95% CI, 0.07-0.33]) and volume (β=0.21 [95% CI, 0.09-0.34]; Figure 2). Among subtypes, hypertension was associated with deep lacune count (β=0.16 [95% CI, 0.04-0.29]) and volume (β=0.19 [95% CI, 0.06-0.31]), but not with lobar lacune measures. HDL cholesterol, current smoking status, and diabetes mellitus were not significantly associated with any lacune measure. Associations of APOE ε4 status with any lacune measure were not statistically significant.

Compared to White participants, Hispanic participants had lower overall lacune count (β=−0.27 [95% CI, −0.45 to −0.09]) and volume (β=−0.29 [95% CI, −0.46 to −0.11]; Figure 2). Deep lacune measures in Hispanic participants trended lower, consistent with their reduced overall lacune burden, but did not survive correction for multiple comparisons. Neither Black nor Chinese American participants differed significantly from White participants on any lacune outcome

### 3.4. Association of Lacune Burden with Cognition and Neuroimaging Markers

Overall lacune count showed significant associations with hippocampal volume, total WMH, periventricular WMH, subcortical WMH, total ePVS, basal ganglia ePVS, and thalamic ePVS, most strongly with subcortical WMH (β=0.14 [95% CI, 0.08-0.20]) and thalamic ePVS (β=0.13 [95% CI, 0.07-0.19]; Figure 3). Overall lacune volume was significantly associated with total WMH (β=0.15 [95% CI, 0.09-0.20]), periventricular WMH, subcortical WMH, basal ganglia ePVS, and thalamic ePVS. Overall lacune measures were not significantly associated with cognitive measures, SPARE-AD, SPARE-BA, cortical gray matter volume, or CMB counts after multiple comparison correction.

**Figure 3.**
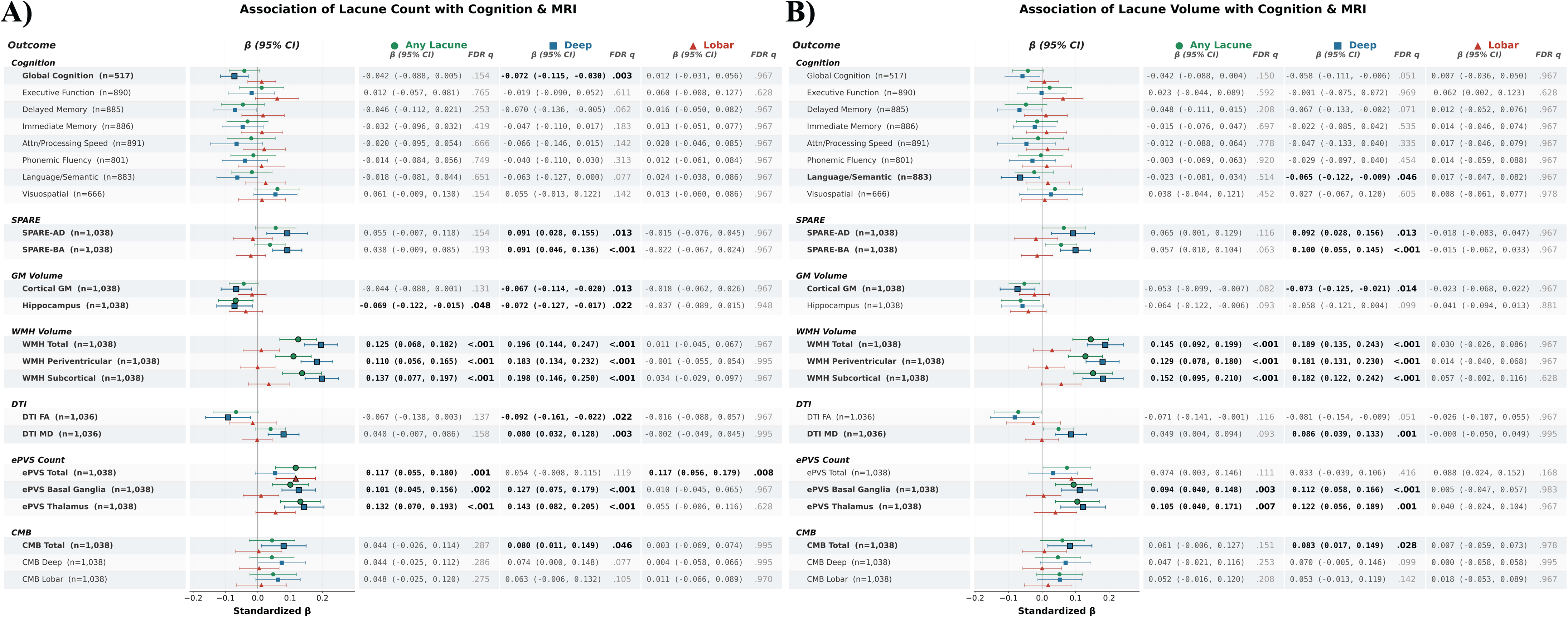
Association of Lacune Burden With Cognition and Neuroimaging Markers. Forest plots showing associations of lacune count (A) and volume (B) with cognitive and neuroimaging outcomes, stratified by lacune subtype. Point estimates represent β coefficients with 95% CIs. Bold rows indicate FDR q < 0.05. Abbreviations: CI, confidence interval; CMB, cerebral microbleed; DTI, diffusion tensor imaging; ePVS, enlarged perivascular space; FA, fractional anisotropy; FDR, false discovery rate; GM, gray matter; MD, mean diffusivity; SD, standard deviation; SPARE-AD, Spatial PAttern REcognition of Alzheimer Disease; SPARE-BA, Spatial PAttern REcognition of Brain Aging; WMH, white matter hyperintensity.

Deep lacune burden showed the most widespread associations with neuroimaging and cognitive markers (Figure 3). Deep lacune count was associated with decreased global cognition (β=−0.07 [95% CI, −0.12 to −0.03]), higher SPARE-AD (β=0.09 [95% CI, 0.03-0.15]) and SPARE-BA (β=0.09 [95% CI, 0.05-0.14]), lower cortical gray matter and hippocampal volumes, greater total, periventricular, and subcortical WMH (β=0.20 [95% CI, 0.15-0.25] for subcortical WMH), lower DTI FA, higher DTI MD, greater basal ganglia and thalamic ePVS, and higher total CMB count.

Deep lacune volume showed parallel associations with decreased language/semantic performance (β=−0.07 [95% CI, −0.12 to −0.01]), higher SPARE-AD (β=0.09 [95% CI, 0.03-0.16]) and SPARE-BA (β=0.10 [95% CI, 0.06-0.14]), lower cortical gray matter, greater total, periventricular, and subcortical WMH (β=0.19 [95% CI, 0.13-0.24] for total WMH), higher DTI MD, greater basal ganglia and thalamic ePVS, and higher total CMB count. Global cognition and delayed memory showed nominally significant associations with deep lacune volume that did not survive multiple comparison correction.

Lobar lacune burden was not significantly associated with cognitive measures, SPARE-AD, SPARE-BA, gray matter volumes, WMH measures, or CMB measures after multiple comparison correction (Figure 3). Lobar lacune count was associated only with total ePVS (β=0.12 [95% CI, 0.06-0.18]). Lobar lacune volume showed nominally significant associations with total ePVS and executive function that did not survive multiple comparison correction.

### 3.5. Association of Lacunes with Longitudinal Cognitive Decline

Deep lacune volume was significantly associated with accelerated decline in executive function (β_Lacune×time_ = −0.023 [95% CI, −0.038 to −0.008]; Figure 4). Total lacune volume also showed an association with accelerated executive function decline (β_Lacune×time_ = −0.023 [95% CI, −0.038 to −0.008]). In analyses modeling lacune volume independent of count, deep lacune volume remained significantly associated with increased rate of executive function decline (β_Lacune×time_ = −0.024 [95% CI, −0.039 to −0.009]), and total lacune volume continued to predict accelerated executive function decline, indicating that lacune volume contributes to longitudinal cognitive decline beyond what is captured by lacune count alone. Lobar lacunes were not significantly associated with longitudinal decline in any cognitive domain. In sensitivity analysis, deep lacune volume was associated with accelerated executive function decline independent of other cSVD lesions (β_Lacune×time_ = −0.023 [95% CI, −0.037 to −0.008]; Figure S3). Main effects of lacunes and time are presented in Tables S5 and S6.

**Figure 4.**
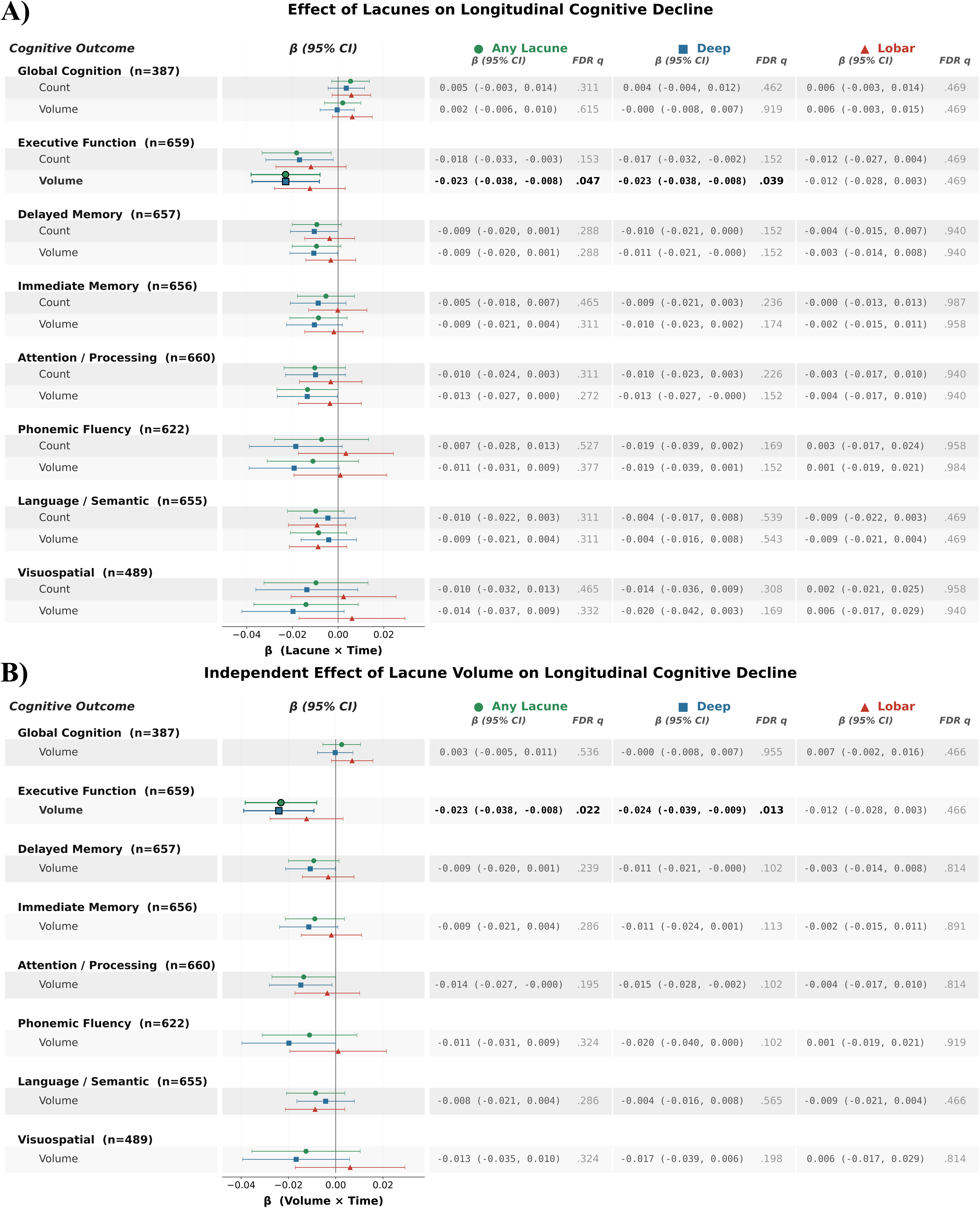
Association of Lacune Burden and Independent Effect of Lacune Volume on Longitudinal Cognitive Decline. Forest plot showing lacune×time interaction coefficients from linear mixed-effects models testing whether baseline lacune burden (count, top row within each domain; volume, bottom row) predicts rate of change in cognitive performance compared to follow-up. Point estimates represent lacune×time interaction β coefficients with 95% CIs. Bold rows indicate FDR q < 0.05 for the interaction term. Abbreviations: CI, confidence interval; FDR, false discovery rate; SD, standard deviation.

### 3.6. Mediation of Age and the Framingham All-CVD Risk Score with SPARE-AD, SPARE-BA, and Global Cognition by Deep Lacunes

In mediation analyses, deep lacune count statistically significantly accounted for a portion of the association between age and SPARE-AD (3.0%), SPARE-BA (1.6%), and global cognition (3.0%; Figure 5). Deep lacune volume showed a similar pattern for age, with a substantially greater proportion of the total association mediated for SPARE-AD (2.8%) than for SPARE-BA (1.6%).

**Figure 5.**
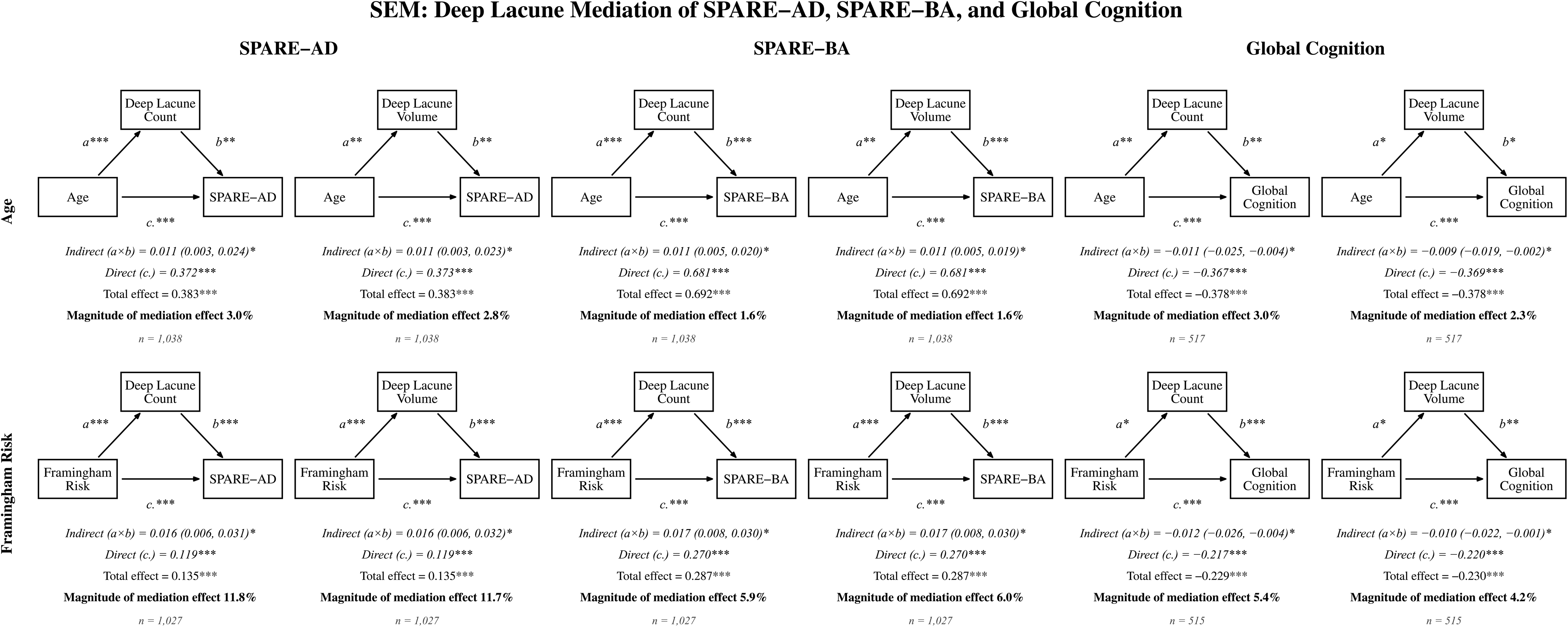
Mediation of Age and Framingham Risk Effects on Cerebral Atrophy and Global Cognition by Deep Lacune Burden. Path diagrams from structural equation models testing whether deep lacune burden mediates the associations between age (top row) or Framingham All-CVD Risk Score (bottom row) and AD-related brain atrophy (SPARE-AD, left column), aging-related brain atrophy (SPARE-BA, middle column), or global cognition (right column). Asterisks on paths a, b, and c′ denote significance of the individual path coefficients (*p<0.05, **p<0.01, ***p<0.001). The indirect (mediation) effect is marked with an asterisk when its bias-corrected bootstrap 95% CI (5,000 resamples) excludes zero. Abbreviations: SEM, structural equation model; SPARE-AD, Spatial PAttern REcognition of Alzheimer Disease; SPARE-BA, Spatial PAttern REcognition of Brain Aging.

A comparable pattern was observed for the Framingham All-CVD Risk Score, with a notably larger proportion of the association attributable to the indirect path through deep lacunes than for age. Deep lacune count accounted for a statistically significant portion of the Framingham-SPARE-AD (11.8%), Framingham-SPARE-BA (5.9%), and Framingham-global cognition (5.4%) associations, again with a greater proportion for SPARE-AD than SPARE-BA. Deep lacune volume showed a similar pattern for the Framingham associations (SPARE-AD, 11.7%; SPARE-BA, 6.0%; global cognition, 4.2%).

## 4. Discussion

To our knowledge, this is among the first applications of deep learning to quantify lacune volume as a continuous measure, and to do so across a prospectively enrolled four-group multi-ethnic, community-representing cohort. Because MESA samples community-dwelling adults rather than stroke patients, these estimates better reflect the subclinical lacune burden of the general population. The inclusion of White, Black, Hispanic, and Chinese American participants further allowed associations to be examined in groups historically underrepresented in lacune neuroimaging.

Overall lacunes and deep lacunes were associated with cardiovascular risk as measured by the Framingham All-CVD Risk Score and the PREVENT 10yr Risk Score. Overall lacunes and deep lacunes were also associated with hypertension. Interestingly, lobar lacunes showed no associations with cardiovascular risk factors. Previous work has suggested that deep lacunes are predominantly associated with hypertensive arteriolopathy, while lobar lacunes were characterized in the setting of cerebral amyloid angiopathy^18,19^. APOE ε4 is a known risk factor for CAA, but its lack of association with lobar lacunes in our study likely relates to incomplete genetic penetrance, competing mechanisms for lobar lacunes, and variations in APOE ε4 associations by race and ethnicity^46^. Potential alternative mechanisms for lobar lacunes include large artery atheroembolism, or subclinical cardioembolic events, although at enrollment symptomatic cardiovascular disease was an exclusion. Further research is needed to investigate whether lobar lacunes in community-dwelling and diverse populations are associated with CAA, have a weaker connection with cardiovascular disease than deep lacunes, or arise from a separate pathophysiological pathway.

Despite a higher cardiovascular risk burden, Hispanic participants had lower lacune burden than White participants, a finding that warrants further investigation. Age was associated with increased lacune burden, and age-specific prevalence in MESA generally aligned with previous large epidemiologic studies^4^. Sex was not significantly associated with lacunes, reflecting a lack of consensus in the literature^4,12^.

Deep lacunes were strongly associated with both WMH volume and with basal ganglia and thalamic ePVSs, suggesting that these markers of cerebral small vessel disease may co-occur along the aging-to-vascular-to-atrophy pathway. Deep lacunes were also associated with multiple patterns of brain atrophy, including SPARE-BA and, notably, SPARE-AD. This association with AD-like atrophy was supported by an independent association between deep lacunes and decreased hippocampal volume and cortical gray matter volume. Mediation analyses further demonstrated that deep lacunes partially accounted for the associations of aging and cardiovascular burden with both SPARE-BA and SPARE-AD, consistent with prior findings demonstrating similar associations and mediation patterns for WMH. These findings implicate deep lacunes not only as markers of cerebrovascular disease but as contributors to atrophy patterns that overlap with those seen in AD and are consistent with a model in which multiple cSVD features converge on shared patterns of brain atrophy and cognitive decline.

Deep lacunes were cross-sectionally associated with global cognition and language/semantic performance, while deep lacune burden was associated with accelerated executive function decline over time. This pattern may reflect distinct cognitive correlates of subcortical vascular injury, whereby deep lacunes may disrupt thalamocortical circuits underlying broader cognitive function, while cumulative lacune burden may render the brain more vulnerable to progressive executive dysfunction. The longitudinal association between lacune burden and executive decline is consistent with prior literature describing white matter network disruption as a driver for the executive dysfunction seen in aging and cSVD^47^. The cross-sectional associations with performance in language/semantic fluency are novel findings that warrant further investigation, although semantic fluency can also be affected by executive dysfunction^48^. Importantly, deep lacune volume was associated with executive function decline independently of deep lacune count, a confirmation of the importance of lacune volume to fully uncover the pathophysiology and downstream effects of lacunes. Furthermore, deep lacune volume retained significant effect sizes after adjusting for other cSVD lesions, indicating that deep lacunes are an independent contributor to future cognitive decline. Mediation analyses further demonstrated that deep lacunes partially accounted for the association of aging and cardiovascular burden with global cognition. The proportion of the age association accounted for by deep lacunes was modest, indicating that while deep lacunes represent a statistically significant pathway linking age to cognition, the direct association of age with cognition remained dominant. In contrast, the proportion accounted for was consistently higher for the Framingham All-CVD Risk Score than for age, suggesting that deep lacunes capture more of the association of modifiable cardiovascular risk with cognition than they do of age-related associations.

Lobar lacunes, in contrast, did not show any major findings with cognition. Furthermore, compared to deep lacunes, lobar lacunes displayed only modest associations with other cSVD lesions; lobar lacunes were only associated with total ePVS. Of note, similar to lobar lacunes, severely enlarged ePVS in the centrum semiovale have also been linked to CAA. However, the prevalence of CAA in this population is likely low relative to arteriolosclerotic cSVD. Combined with the risk-factor findings, these results suggest further research is needed to establish whether lobar lacunes belong within the cSVD spectrum, represent a separate pathological pathway, or are mixed-etiology lesions.

The study has limitations. First, MRI scans were acquired temporally distant from cognitive assessment. While we adjusted for this interval, residual confounding from the time lag may have attenuated lacune-cognition associations. Second, the low prevalence of lacunes limited statistical power for our analysis, especially when stratifying by subtypes. Third, because deep lacune burden and the atrophy outcomes were assessed concurrently on a single MRI, the mediation models cannot establish temporal precedence and warrant confirmation in longitudinal studies. Additionally, our model only achieved a precision of 0.41 and required manual reader quality control. While this partially represents the difficulty of automated lacune detection^49,50^, more work is needed for an independent quantification model. Finally, future work should include larger lacune training sets.

## 5. Conclusion

The present study introduces a deep learning framework for lacune detection and volumetric quantification in a large, multi-ethnic cohort. By enabling automated segmentation, this approach allowed us to quantify lacune volume, a metric previously unavailable in manual rating studies. Deep lacune burden demonstrated stronger associations with vascular risk, other cerebral small vessel disease markers, brain atrophy, and longitudinal executive-function decline than lobar burden. Lacune volume provided information beyond lacune count.

## Non-standard Abbreviations and Acronyms

CMB: cerebral microbleed
cSVD: cerebral small vessel disease
CVD: cardiovascular disease
DTI: diffusion tensor imaging
ePVS: enlarged perivascular space
FA: fractional anisotropy
FDR: false discovery rate
FLAIR: fluid-attenuated inversion recovery
ICV: intracranial volume
MARS: Microbleed Anatomical Rating Scale
MD: mean diffusivity
MESA: Multi-Ethnic Study of Atherosclerosis
MUSE: MUlti-atlas region Segmentation utilizing Ensembles
SPARE-AD: Spatial PAttern REcognition of Alzheimer’s Disease
SPARE-BA: Spatial PAttern REcognition of Brain Aging
STRIVE: STandards for ReportIng Vascular changes on nEuroimaging
WMH: white matter hyperintensity

## 6. Acknowledgments

We thank the other investigators, the staff, and the participants of the Multi-Ethnic Study of Atherosclerosis (MESA) study for their valuable contributions. A complete list of participating MESA investigators and institutions can be found at http://www.mesa-nhlbi.org.

## 9. Data Availability

MESA data are available through the NHLBI BioLINCC repository (https://biolincc.nhlbi.nih.gov), with genetic data available through dbGaP, upon application and approval. The lacune deep learning model and QC tool is available at https://github.com/UTHSCSA-NAL.

## 10. Supplemental Material

Supplemental Methods

Lacune QC SOP

Supplemental Figures

Figures S1-S3

Tables S1-S6

